# Friday excess of overall mortality in three German subpopulations

**DOI:** 10.1101/2023.09.12.23295427

**Authors:** Jiří Wackermann

## Abstract

We studied variations of all-cause human mortality during the weekly cycle, using deathdates collected from death notices published in regional or local newspapers in three distinct regions in Germany (South Baden, North Bavaria, Saarland), total database size *N* ≈ 44000. Probability distributions of death on different days of the week show significant (*P* = 0.0155) departures from the uniformity hypothesis. The day of maximum mortality is Friday (probability of dying = 0.148), whereas minimum mortality is observed on Sundays and Mondays (probability of dying = 0.140 for both days). This ‘Friday excess’ effect seems to be a novel phenomenon, unrelated to other findings on weekly modulation of human mortality.

## Introduction

Study of factors determining and modulating human mortality are of eminent importance for medical and social sciences. Among these, temporal factors are of particular interest. While monotonically increasing as a function of age, mortality exhibits periodic variations during the annual ^1^–3 and diurnal ^4^,5 cycle. There is also a sizeable literature on weekly variations of cause-specific mortality, e. g. due to cardiovascular diseases ^6^–11. Less explored is the weekly course of mortality in general, non-clinical populations. Here we report a remarkable pattern of weekly variations in all-cause mortality observed in three geographically distinct subpopulations in Germany.

## Materials and Methods

Data were collected from death notices publicly available via web portals of local or regional, weekly or daily newspapers. Decedents’ birthdates and deathdates, gender, and names were excerpted. Only death cases with these data completely and unequivocally specified were included into the database. Four datasets were created, distinguished by data sources: **wzo** ^12^ (merger of five local weekly newspapers, district of Emmendingen), **bz** ^13^ (*Badische Zeitung*, daily, South Baden), **nba** ^14^ (merger of several daily local newspapers, North Bavaria), **saa** ^15^ (*Saarbrücker Zeitung*, daily newspaper, Saarland). Datasets were revised for names/dates conflicts and multiple references to decedents were eliminated. Finally, deaths expressly due to accidents or ‘tragic’ circumstances, or at the decedent’s age < 40 years, were removed. Detailed descriptions of the datasets are given in Table 1.

**Table 1.** Datasets description.

| Source ID | Observation epoch |  |  | Number of cases |  |  | Mean age (SD) |  |
| --- | --- | --- | --- | --- | --- | --- | --- | --- |
|  | from | to | days | men | women | total | men | women |
| **wzo** | 1 Jan 2016 | 31 Dec 2022 | 2557 | 2549 | 2490 | 5039 | 78.1 (11.5) | 83.1 (11.4) |
| **bz** | 1 Jan 2018 | 31 Dec 2019 | 730 | 5847 | 5945 | 11792 | 79.3 (11.1) | 83.7 (11.0) |
| **nba** | 1 Jan 2018 | 31 Dec 2019 | 730 | 9126 | 9248 | 18374 | 78.6 (11.2) | 83.1 (10.7) |
| **saa** | 1 Jan 2018 | 31 Dec 2019 | 730 | 7034 | 7244 | 14278 | 78.9 (10.8) | 83.1 (10.6) |

Deathdates were transformed to indices of days of the week ^16^ (Monday → 1, …, Sunday → 7) and numbers of death cases *n*_*k*_ falling on different days of the week (*k* = 1, …, 7) were evaluated. Relative frequencies 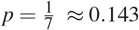 (where *N* = ∑_*k*_ *n*_*k*_ = total dataset size) provide estimates of probability of dying on the day *k*. The constant mortality hypothesis implies a uniform distribution of deaths throughout the week, 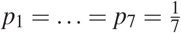.^†^ To test the uniformity hypothesis we use Pearson’s statistics

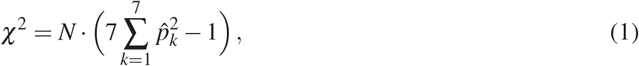

which would be chi-square distributed with *df* = 6. To evaluate qualitative resemblance of two vectors of probabilities *p*, we use a measure of dissimilarity defined by a sum of absolute rank differences

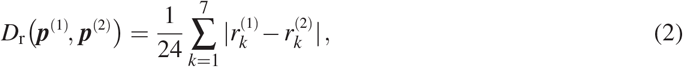

where *r*_1_, …, *r*_7_ denote ranks of elements *p*_1_, …, *p*_7_ of the vectors *p*. Under uniformity assumption, all permutations of ranks 1, …, 7 are equally probable, so that the *P* value corresponding to an observed *D*_r_ value can be calculated by complete enumeration.

## Results

First time we observed the effect of our interest in a small dataset **wzo**, consisting of about five thousand death cases collected from five local newspapers over a period of seven years. We noticed remarkable variations of mortality across the week, namely, an excess of deaths on Fridays and a deficit thereof on Sundays and Mondays (Fig. 1A). Even if the deviations from the uniform distribution are not statistically significant (χ^2^ = 6.239, *df* = 6, *P* = 0.397), this observation motivates a closer look at the phenomenon. Therefore we analysed three large datasets **bz**, **nba** and **saa** collected from regional newspapers during years 2018 and 2019. There (Fig. 1B) we find patterns clearly resembling those seen in the dataset wzo (Fig. 1A). Again, deviations from the uniform distribution are not large enough to qualify as ‘significant’ (**bz** : χ^2^ = 2.717, *P* = 0.843; nba : χ^2^ = 10.837, *P* = 0.093; **saa** : χ^2^ = 5.522, *P* = 0.479; all *df* = 6). However, the weekly courses of relative frequencies show notable commonalities in terms of their ranks—**bz**: 3461752, nba: 1435762, **saa**: 3451762. (Reading from the left to the right in the order Monday to Sunday). In particular, the highest mortality falls without exception on Friday.

**Figure 1.**
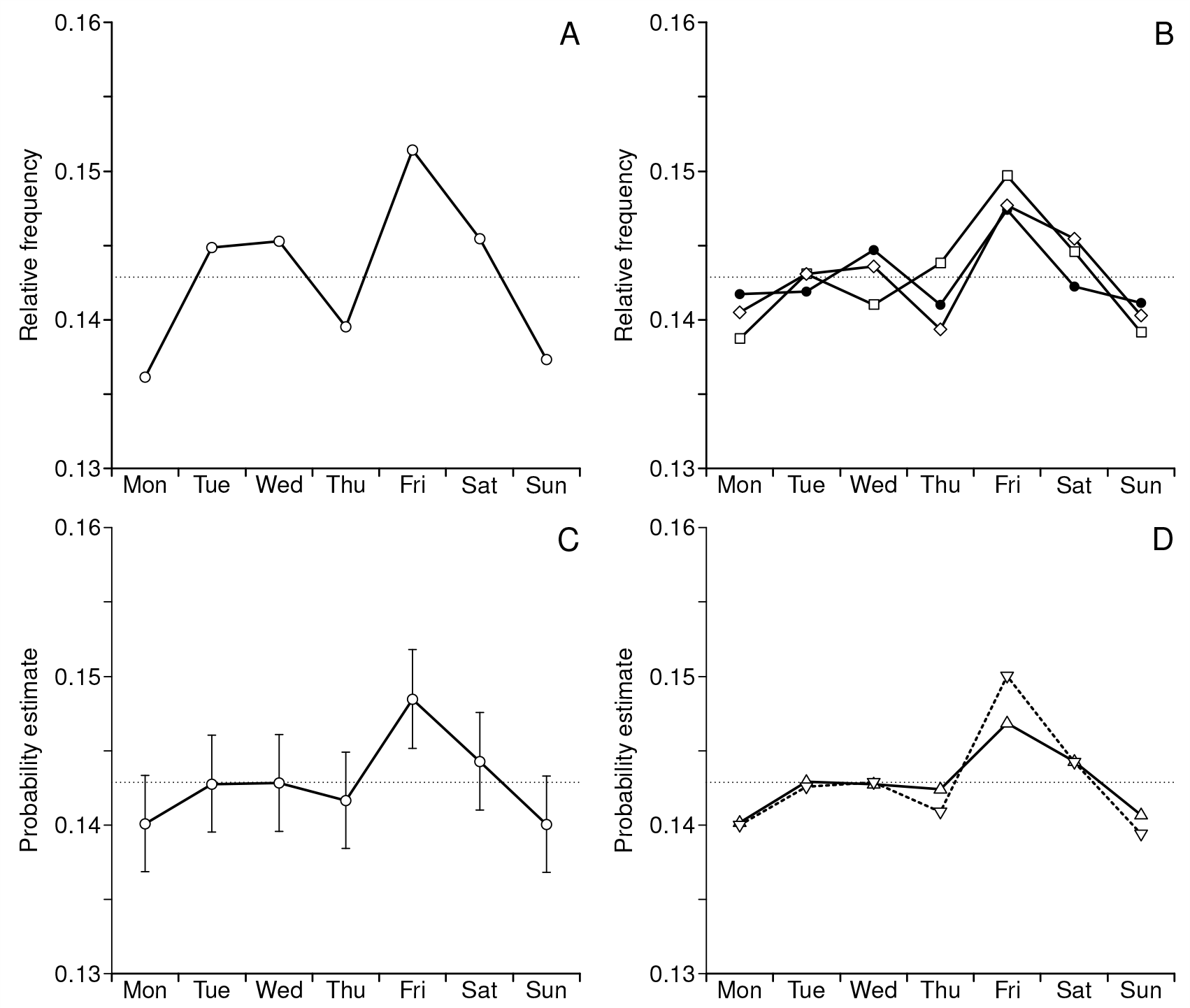
Relative frequencies of death as functions of the day of the week. A: dataset **wzo**. **B**: three separate datasets **bz** (•), **nba** (□) and **saa** (◊). C: Combined dataset **com**; estimates of death probabilities shown along with 95% CIs. D: Subsets **comM** (men, Δ, solid line), **comF** (women, ▽, dashed line). In all four panels the dotted horizontal line indicates uniform distribution 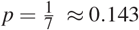.

Next, we combine the three datasets into a superset **com** = **bz** ∪ **nba** ∪ **saa** comprising, after removal of a few duplicities, a total of 44428 death cases. Here the weekly course of death frequencies (Fig. 1C) shows the common pattern even more conspicuously. The test of uniformity yields χ^2^ = 15.684, *df* = 6, *P* = 0.0155, so we now safely reject the uniformity hypothesis. The similarity between patterns seen in datasets **wzo** and **com** is obvious: their rank-transformed sequences are almost identical (com: 2453761, wzo: 1453762, *D*_r_ = 0.083) except for the two lowest values on Sunday and Monday. Evaluating 95% confidence intervals for the mortality estimates, we find 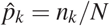, CI = [0.145, 0.152], that is, a significantly higher probability of death on Friday. The relative risk, i. e. the ratio of Friday vs. mean non-Friday mortality, is then 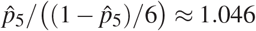.

Sex-related differences in the magnitude of the effect described may be also of interest. Fig. 1D shows the weekly distributions of deaths for two subsets of the combined dataset, **comM** (men, *N*_M_ = 21998) and **comF** (women, *N*_F_ = 22430). χ^2^-tests indicate only marginally significant (women, χ^2^ = 12.133, *P* = 0.059) or none (men, χ^2^ = 4.658, *P* = 0.588) deviations from uniformity. The two sex-specific mortality patterns are very similar, whether quantified by χ^2^ statistics (2 × 7 contingency table, χ^2^ = 1.082, *df* = 6, *P* = 0.982) or in terms of ranks (comM: 1543762, comF: 2453761, *D*_r_ = 0.167).

## Discussion

This is, to the best of our knowledge, the first observation of the particular weekly pattern of all-cause mortality characterised by a significant excess of Friday mortality along with its noticeable decrease on Sundays and Mondays. Note that we use the expression ‘Friday excess’ as a shortcut for the Friday– Sunday–Monday pattern in its entirety. The effect is very small and requires a dataset of considerable size to be detected. This may explain why the effect has escaped researchers’ attention until now. We find this pattern expressed significantly (*P* = 0.0155) in a dataset combined of three datasets of different origin, **bz**, **nba**, and **saa**, comprising about forty thousands death cases, but not for separate datasets. In our view, however, the consistent pattern of mortality variation occurring across the three subsets—i. e. across three disjoint subpopulations—weights no less than conventional measures of significance.

Interpretation of the ‘Friday excess’ effect remains a challenging task. We may try to explain away the phenomenon as an artefact emerging during the data reporting, coding or processing phase. This, however, meets serious difficulties. Data for death notices are submitted to the newspapers by persons from the decedents’ social environment, most frequently their families. A mechanism of biased reporting or publication filtering is beyond all imagination. Days of the week are derived from death dates on our side of the data processing chain, so there is no place for systematic errors to take place.

Assuming the phenomenon is real, we have to look for possible psychosocial or physiological backgrounds. As mentioned in the introduction, there are numerous findings regarding weekly variations of mortality but their relevance for our study is doubtful. We leave aside reports regarding effects of the day of the week of a medical intervention (e. g. hospital admission ^17^ or surgical treatment ^18^) on patients’ mortality, which are presumably unrelated to the effect reported here. Of some interest may be variations of cause-specific mortality during the week cycle, such as the well-known increase of frequency of deaths from cardiovascular events usually associated with Monday ^6^–9, though not without exceptions ^10^. Another facet of the ‘Monday effect’ seems to be the maximum frequency of suicides observed on Mondays in contrast to lowest suicide numbers on weekends ^19^–21. Dynamics of positive and negative affects was shown to follow the seven days cycle even in a clinically healthy sample, again with a maximum of negative affect on Mondays ^22^. We may also mention a study of all-cause mortality in Jewish population ^23^ showing a decrease/increase pattern on Saturdays/Sundays, where Saturday as the day of rest corresponds to Sunday in Western culture.

A common feature of these findings is the prominence of Monday as the ‘critical’ day, marking the increased workload and stress on weekdays and thus possibly related to rising mortality ^6^,11. In case of the ‘Friday effect’, however, we have a reversed pattern: we observe a significant increase in mortality on Fridays, i. e. the days preceding the period of rest, followed by a mortality decrease which extends up to Monday. The ‘Monday effects’ cited above and the ‘Friday excess’ effect described in this paper are not easy to reconcile.

The present study certainly has some weaknesses, especially the lack of information on individual death causes and a rather vague delimitation of the populations under study. Further studies of the effect based on data from general but more precisely characterised populations are definitely desirable. The aim of this brief report is just to communicate a novel phenomenon in human mortality and to encourage its further exploration.

## Data Availability

All data produced in the present study are available upon reasonable request to the authors.

## Acknowledgements

Thanks are due to Anna Yamamotová and Werner Ehm for their reading and commenting on an earlier version of the paper.

## Footnotes

† To be correct, this holds exactly only for balanced numbers of days of the week in the data series. Otherwise the expected frequencies of deaths should be adjusted to to actual numbers of days of the week within the observation epoch. However, post hoc calculations reveal that effects of unbalanced numbers of days of the week are small and do not affect our results essentially.

